# Neurodivergence as a risk factor for Post-Covid-19 Syndrome

**DOI:** 10.1101/2023.06.08.23291154

**Authors:** Rachael K. Raw, Jon Rees, Amy Pearson, David R. Chadwick

**Affiliations:** School of Medicine and Health, Newcastle University, Newcastle upon Tyne, UK; The School of Psychology, University of Sunderland, UK; Centre for Clinical Infection, James Cook University Hospital, Middlesbrough, UK

**Keywords:** SARS-CoV-2, COVID-19, Long-COVID, Post-Covid-10 Syndrome, Chronic Fatigue Syndrome, Autism, Neurodivergence, Neurotypical

## Abstract

Neurodivergent (ND) individuals (e.g., Autistic people) are more likely to experience health problems that are characterised by central sensitisation ’. Recent research suggests that a so-called ‘Long-COVID’ syndrome might also be explained by a heightened response to internal physiological stimuli, much like in myalgic encephalomyelitis/chronic fatigue syndrome (ME/CFS). Using a standardised assessment tool, we examined whether traits associated with Autism would predict long-term COVID-19 symptoms in 267 Healthcare Workers (HCW).. Higher autistic traits predicted COVID-19 symptoms that lasting more than 12 weeks regardless of formal autism diagnosis. A personality measure also showed that negative affect was associated with experiencing COVID-19 symptoms for 4-12 weeks, though the direction of causality in this case is uncertain. Limitations of the present study are 1) the retrospective nature of COVID-19 symptom reporting; 2) likely self-selection bias given the high number of HCWs who reported long-term COVID-19 symptoms; and 3) the gender-bias towards females in our sample.

## Introduction

Since the onset of the global COVID-19 pandemic in 2020, researchers have sought to understand the long-term consequences of SARS-CoV-2 infection. Guidelines have been published in attempts to define a ‘Long-COVID’ syndrome, suggesting that the duration of COVID-19 symptoms can be categorised as either acute COVID-19 Symptoms (AC) lasting up to 4wks post-infection; ongoing symptomatic COVID-19 (OSC) with symptoms up to 12wks; or post-COVID-19 syndrome (PCS); when symptoms span beyond 12 weeks – more commonly known as Long-COVID^[1]^. Long-term symptoms can include physiological sequalae, typically shortness of breath (SOB), cough, headache, myalgia/arthralgia, fatigue and/or psychological outcomes such as cognitive impairment, memory loss, anxiety and depression^[2]^. Recent studies suggest that some individuals are more prone to developing long-term symptoms of COVID-19, with females, smokers and those with pre-existing co- morbidities being the most at-risk^[3-8]^. Furthermore, symptoms of Long-COVID have been likened to those that characterise myalgic encephalomyelitis/chronic fatigue syndrome (ME/CFS) - a chronic multi-system condition, that has been found to be triggered by infectious diseases such as Influenza and Coronaviruses^[8,9]^.

While the mechanisms underlying ME/CFS are not fully understood, a key theory is that of ‘Central Sensitization’ (CS), whereby the nervous system has an amplified response to certain stimuli. For example, a heightened reaction to nociceptive input may lead to hyperalgesia and thus a heightened experience of pain^[10]^. This theory has been applied to certain groups, where an amplified internal response to external stimuli is commonplace, such as neurodivergent (ND) people (e.g. Autistic people). Accordingly, a recent study found that a large proportion of autistic individuals met the diagnostic criteria for ME/CFS, as well other conditions characterised by CS, including fibromyalgia and irritable bowel syndrome (IBS). Sixty-percent of autistic participants also scored at or above the clinical cut-off on a screening tool for CS^[10]^. Considering this evidence, we aimed to examine whether autistic individuals, and those with higher self-reported autistic characteristics are also at greater risk of developing long-term symptoms of COVID-19. As some evidence suggests that Type D traits, including negative affect and social inhibition, are more common in those with conditions such as ME/CFS and fibromyalgia,^[11]^ this measure was included to control for these as a possible confounder. In the present study, we therefore surveyed a group of healthcare workers (HCWs) and asked them about their experience of ongoing COVID-19 symptoms. Our main aim was to see whether longer-term COVID-19 symptoms were more common in those who scored higher on a measure of autistic characteristics in ND and Type D Personality (TDP).

## Method

### Ethics Statement

All participants gave their written informed consent, and Cambridge East Research Ethics Committee approved this study (Ethics Ref: 20/EE/0161), which was performed in accordance with the ethical standards laid down in the 1964 Declaration of Helsinki. Data was collected between August and November 2022. Authors were blinded to identifiable participant information.

### Participants

In this observational survey-based study, HCWs were recruited from three Northeast England hospitals and formed an opportunistic sample. HCWs were invited via email to complete an online survey, which was advertised to all HCWs using the weekly communications service.

### Materials and Procedure

The anonymous survey explored HCWs’ experience of COVID-19 symptoms, including nature, severity and duration. Prior COVID-19 history was defined as previous positive self-reported polymerase chain reaction (PCR), antibody (AB) and/or lateral flow test (LFT). HCWs indicated which COVID-19 symptoms they experienced from the following list: chest pain; memory/concentration problems; cough; diarrhoea; fatigue; fever/chills; headache; joint pain; loss taste/sell; muscle pain; nausea/vomiting; SOB; sore throat; swollen lymph nodes. Severity was ranked as ‘Mild’ (no interference with daily activities); ‘Moderate’ (some interference); ‘Severe’ (prevented daily activities); ‘Very Severe’ (required hospital visit/hospitalisation). Symptoms duration was categorised according to previously cited guidelines^[1]^, as AC, OCS, or PCS. Vaccination history was also recorded. HCWs were invited to self-declare whether they were neurodivergent. Autistic traits were assessed using the Ritvo Autism & Asperger Diagnostic Scale (RAADS-14)^[12]^. TDP was explored using the DS14, a psychometric tool that captures negative affectivity and social inhibition^[13]^. Effects of age and gender were also considered within our analyses.

### Statistical Analysis

Statistical analysis was conducted using JASPv0.16.3.0^[14]^. Categorical outcomes were examined with chi-square tests and multinomial regression. Continuous outcomes were investigated using ANOVA. An alpha level of .05 indicating significance was used throughout.

## Results

Of the 267 respondents (35 M, 226 F, 6 other; mean age 48.3yrs), 153 (57%) reported one episode of COVID-19, 100 (37%) reported two episodes, and 14 (5%) recalled three or more, which had been diagnosed on positive PCR (208, 78%), LFT (158, 41%) or AB test (33, 12%) with 111 positive on both PCR and LFT. 212 (79.5%) received three doses of vaccine, 23 (8.5%) more than three doses, 24 (9%) had two doses, 3 (1%) one dose and 3 (1%) none. Two (1%) declined to give vaccine information. Regarding COIVD-19 symptoms 124 (46%) resolved within 4wks (AC), 64 (24%) between 4-12wks (OCS) and 79 (30%) had symptoms >12wks (PCS).

Thirty-eight (14%) self-declared being ND (16 autistic; 14 attention deficit hyperactivity disorder; 21 others including dyslexia, dyscalculia, dyspraxia; 12 with multiple conditions), whilst 7 (3%) declined to answer. There was no significant association between either OCS or PCS and self-declared ND status (Χ^2^(2)=2.59, p=.27). Based on the RAADS cut-off of 14, 83 (31%) scored above this threshold and there was a significant association of being above threshold with likelihood of experiencing OCxS or PCS (Χ^2^(2)=10.01, p=.007).

There was a significant association between number of COVID-19 episodes and symptom duration (Χ^2^(4)=13.57,*p*=.009) with 2 episodes being associated with PCS. Increased symptom number during first episode was reporting by people going on to develop either OCS or PSC (7.5(2.9) or 8.3(3.0) vs 5.1(2.9) symptoms; *F*(2,272)=34.46, *p*<.001), with the increased occurrence of every symptom being associated with OCS or PCS except fever and sore throat. Those reporting PCS also reported greater severity of symptoms during the first episode compared to those with OCS, or those whose symptoms resolved (Χ^2^(6)=43.90, *p*<.001). There was no association of reported first episode symptom severity with ND status (Χ^2^(6)=2.58, *p*=.86).

Examining the subscales of the RAADS, there was no difference between COVID-19 duration groups on mentalising (*F*(2,259)=2.37, *p*=.10) or social anxiety (*F*(2,259)=2.5, p=0.08), but a significant difference in sensory reactivity (*F*(2,259)=4.85, *p*=.009), with post-hoc testing showing that there was a significant difference between the AC and PCS (1.62 (2.49) vs 3.81 (3.02)). There was no overall difference in DS14 between the COVID-19 duration groups (*F*(2,259)=2.41, p=.09). Negative affectivity was significantly higher in those experiencing OCS (*F*(2,272)=3.72, *p*=.03), but not those with PCS. See Table 1 for HCW characteristics.

**Table 1.**
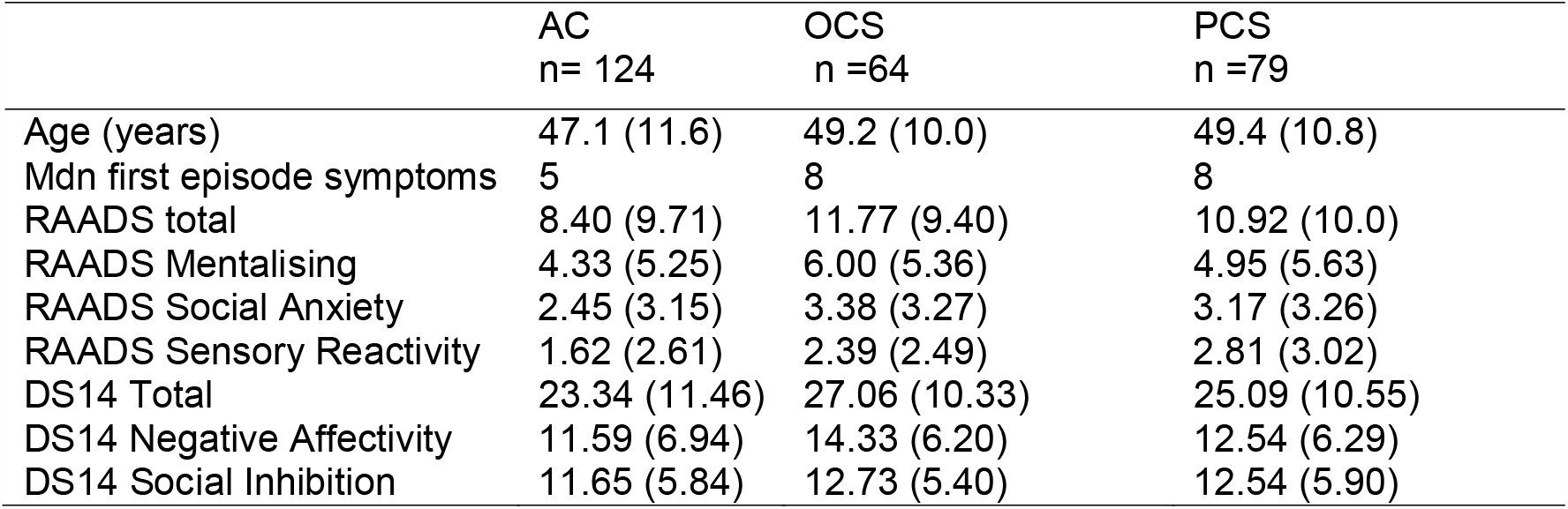
Characteristics of the three patient groups showing mean (SD) for age, totals and subscale totals for the RAADs and DS14 and median number of symptoms reported during first COVID-19 episode.

A multinomial logistic model was constructed with three levels of outcome being symptom duration. Age and gender were included in the model at step one, and the two TDP subscales and three RAADs subscales entered at a second step. The second step resulted in a significant improvement in model fit (Χ^2^(10)=22.75, p=.012). Only the DS14 subscale for negative affectivity was a significant predictor of OCS versus AC. PCS was significantly predicted by the RAADS sensory reactivity subscale as well as increasing age and being female. Coefficients are given in Table 2. Repeating the analysis with OCS symptoms as the baseline, none of the factors significantly differentiate that from the PCS group.

**Table 2.** Coefficients from multinomial logistic regression predicting occurrence of OCS or PCS against baseline AC from the three RAADS and two DS14 subscales.

|  | Estimate (S.E.) | Odds Ratio (95% C.I.) | p |
| --- | --- | --- | --- |
| **OCS v AC** |  |  |  |
| Intercept | -2.99 (1.03) |  |  |
| Age | 0.03 (0.02) | 1.03 (0.99 – 1.06) | .07 |
| Sex | M: -0.37 (1.34) | 0.69 (0.05 – 9.63) | .79 |
|  | F-M 0.62 (0.51) | 1.86 (0.68 – 5.08) | .23 |
| DS14 Negative Affectivity | 0.07 (0.03) | 1.07 (1.01 – 1.15) | .04 |
| DS14 Social Inhibition | -0.08 (0.05) | 0.92 (0.84 – 1.01) | .92 |
| RAADS mentalising | -0.003 (0.05) | 1.00 (0.91 – 1.10) | .95 |
| RAADS Social Anxiety | 0.13 (0.08) | 1.14 (0.98 – 1.33) | .09 |
| RAADS Sensory Reactivity | 0.09 (0.08) | 1.10 (0.93 – 1.29) | .27 |
| **PCS v AC** |  |  |  |
| Intercept | -3.10 (1.02) |  |  |
| Age | 0.03 (0.02) | 1.03 (1.00 – 1.06) | .04 |
| Sex | M: -0.05 (1.31) | 0.96 (0.07 – 12.50) | .97 |
|  | F-M: 1.13 (0.54) | 3.09 (1.07 – 8.88) | .04 |
| DS14 Negative Affectivity | 0.01 (0.03) | 1.01 (0.95 – 1.08) | .70 |
| DS14 Social Inhibition | -0.04 (0.05) | 0.96 (0.88 – 1.05) | .40 |
| RAADS mentalising | -0.07 (0.05) | 0.93 (0.85 – 1.02) | .12 |
| RAADS Social Anxiety | 0.13 (0.08) | 1.14 (0.98 – 1.32) | .09 |
| RAADS Sensory reactivity | 0.22 (0.08) | 1.25 (1.07 – 1.46) | .004 |

A similar model revealed that after controlling for age, gender, RAADs sensory reactivity and TDP negative reactivity, OCS was significantly predicted by the presence of SOB (O.R. 6.14 [2.64–14.29], *p*<.001). PCS was predicted by the occurrence of concentration or memory problems (O.R. 5.59 [2.28–13.72], *p*<.001), during first episode. Fever during the first episode was associated with lower likelihood of OCS (O.R. 0.37 [0.15–0.90], *p*=.03 or LC O.R. 0.21 [0.08–0.56], *p*=.002).

## Discussion

Conditions characterised by CS, such as ME/CFS, are more common in those with ASD^[11]^. PCS, where symptoms of SARS-CoV-2 infection last beyond 12 weeks^[1]^, has been likened to ME/CFS^[8,9]^. We explored whether neurodivergent HCWs were more likely to experience prolonged symptoms of COVID-19 and whether TDP traits are a risk factor for PCS.

In our sample 30% described having COVID-19 symptoms beyond 12wks post-infection. This is alarmingly high, given the extent to which ongoing COVID-19 symptoms can impact daily life. Loss of earnings, dependency on caregivers, and an inability to perform everyday activities are just a few of the consequences of PCS cited in the recent literature^[15]^. Some of the previously highlighted risk factors for prolonged COVID-19 symptoms were replicated in the present study, with females and older individuals being more at risk of PCS. Having had multiple episodes of COVID-19 was also associated with longer length of symptoms, and increased symptom number in the first episode was associated with OCS/PCS.

Regarding Type D Personality, TDP traits overall were not different across the AC/OCS/PCS groups. Importantly, the confounder of negative affect was only marginally associated with OCS, and not at all with PCS. The direction of causality in this finding must be carefully considered, as it is unclear as to whether the presence of OCS might have led to greater negative affect, or if those with naturally greater negative affectivity were more likely to have had symptoms for longer because they perhaps did not take the actions necessary to accelerate their recovery (e.g., returning to regular activities).

Interestingly only 13% of HCW self-declared that they were neurodivergent, whereas 31% of the group scored above threshold on the RAADS. This is consistent with evidence to suggest that people with fewer stereotypical characteristics frequently go un-diagnosed^[16]^. Analyses also revealed that PCS was predicted by the ‘sensory reactivity’ subscale of the RAADS, suggesting that HCWs who scored highly on this element were more likely to have prolonged symptoms. This finding fits with the theory that a CS mechanism may underlie conditions such as ME/CFS^[9]^.

The main limitations of the present study are the likelihood of self-selection bias given that 30% of HWCS had experienced PCS, that we relied on HCWs to recall symptoms of COVID- 19 retrospectively, and that the sample was largely comprised of female participants (85%). Nevertheless, our work provides preliminary evidence to support the sensory reactivity element of neurodivergence as a risk factor for developing Long-COVID. This finding has implications for the provision of treatment services in at-risk groups^[16]^, particularly psychological interventions, which autistic individuals often find difficult to access^[18,19]^.

## Conclusion

Our key findings are first, more HCWs scored above threshold for neurodivergence than those who self-declared having been diagnosed neurodivergent; second, there was no association between OCS/PCS and HCWs self-declared neurodivergent status; third, TDP did not differ or predict OCS/PCS, but the negative affectivity subscale was a relatively weak predictor of being in the OCS group; and finally, scoring above or below the RAADS threshold was predictive of PCS, with the sensory reactivity subscale (in addition to age and female sex) predicting COVID-19 symptoms lasting >12 weeks.

## Data Availability

All data produced in the present study are available upon reasonable request to the authors

## Contributors

RKR/JR/DRC/AP conceived the study and DRC is chief investigator of the CHOIS study. RKR acted as site principal investigator. RKR/JR/DRC contributed to the study protocol, design, and data collection. JR did the statistical analysis. RKR/JR/DRC prepared the manuscript. All authors critically reviewed and approved the final version.

## Acknowledgements

We would like to thank the CHOIS research team, John Rouse and the North East and North Cumbria NIHR for assistance with the survey.

## Data Availability

All data will be made available upon request.

## List of Abbreviations

AB: Antibody
AC: Acute COVID-19
CS: Central Sensitisation
DS14: Type D Personality Scale
HCWs: Healthcare Workers
IBS: Irritable Bowel Syndrome
LFT: Lateral Flow Test
ME/CFS: Myalgic Encephalomyelitis/Chronic Fatigue Syndrome
ND: Neurodivergent/Neurodivergence
OSC: Ongoing Symptomatic COVID-19
PCR: Polymerase Chain Reaction
PCS: Post-COVID-19 Syndrome
RAADS: Ritvo Autism & Asperger Diagnostic Scale
SOB: Shortness of Breath
TDP: Type D Personality

